# Assessing mosquito dynamics and dengue transmission in Foz do Iguaçu, Brazil through an enhanced temperature-dependent mathematical model

**DOI:** 10.1101/2025.01.10.25320311

**Authors:** C. S. Rauh, E. C. Araujo, F. Ganem, R. M. Lana, A. S. Leandro, C. A. Martins, F. C. Coelho, C. T. Codeço, L. S. Bastos, S. T. R. Pinho

## Abstract

Dengue fever is a public health concern that demands efforts to mitigate its impact. We aim to investigate the influence of key parameters temperature-dependent on dengue transmission dynamics in Foz do Iguaçu, a triple border municipality in south Brazil, applying a mathematical model composed by systems of ordinary differential equations. Adjusted model simulation is consistent with the observed data. The effective reproduction number was calculated for detecting changes in dengue transmission over time and to timely detect the beginning of epidemics. Additionally, we explore the potential effects of climate variability on dengue dynamics. Our findings show the importance of vector population dynamics, climate and incidence, contributing to a deeper understanding of dengue transmission dynamics in Foz do Iguaçu and providing a foundation for optimizing intervention strategies, also, in other cities, enhancing our ability to predict and manage dengue outbreaks and supporting public health efforts to control measures.

## 1 Introduction

Dengue is a viral vector-borne disease with a complex transmission dynamic and fast-growing burden, posing challenges for surveillance and control programs worldwide. The dengue virus, classified into four serotypes (DENV1-4) [7], is endemic in tropical and subtropical regions of the world and has its distribution expanding to higher altitudes and latitudes, due to the erosion of climate barriers [29].

The primary dengue vectors, *Aedes aegypti* and *Aedes albopictus* mosquitoes, are highly anthropophilic and well-adapted to urban environments. In Brazil, *Ae. aegypti* reinvaded the country after elimination campaigns in the mid-20^th^ century and is now present in most municipalities. These mosquitoes also transmit Zika and Chikungunya viruses, which have been circulating in Brazil since 2014[16].

Dengue transmission dynamics are inherently linked to climatic factors, as they influence the ecology of its invertebrate vectors. Vector density, often measured as the average number of mosquitoes per person, depends on the availability of breeding sites and suitable climate conditions for survival and reproduction [12, 22]. Additionally, there is also a large amount of literature on the effects of climate on mosquito life cycle and vector capacity. Temperature affects the transmission cycle of dengue, by shortening the extrinsic incubation period [10]. The role of temperature on dengue dynamics [30, 22, 13, 46] is well-recognized, with many countries using seasonal climate data to build predictive models and issue dengue warnings [37, 28, 40, 12, 23]. Mathematical models are essential tools for understanding arbovirus transmission dynamics.

By incorporating both human and vector components and being parameterized with comprehensive datasets, these models provide valuable insights for testing hypotheses about disease transmission and exploring intervention scenarios. Such exploratory simulations can predict future outbreaks, improve preparedness, and support the planning of effective vector control interventions and timely responses — a critical challenge for public health authorities. However, despite this potential, modelers rarely have access to long-term measures of both entomological (adult mosquito indices) and epidemiological time series (incidence) to fit these models and test hypotheses [41, 42, 12]. Foz do Iguaçu, located in southern Brazil, serves as a unique and strategic site for assessing dengue transmission at the fringe of the disease distribution. This city lies in the tri-border area with Argentina (Puerto Iguazu) and Paraguay (Ciudad del Este), making it a critical point for studying transboundary dengue dynamics. The region is characterized by a humid subtropical climate with cool winter and warm summer. Foz do Iguaçu has recorded dengue fever since 1998, with nine outbreaks between 2010 and 2021 according to [5] data. Since 2017, the city has operated an unique integrated entomological and epidemiological surveillance system. The system monitors a large set of adult mosquito traps across the city every two months and detects viruses in mosquitoes and human samples, showing good predictive value [25]. This system detected a drastic increase in mosquito abundance during the 2020-2022 period. This rich dataset provides an opportunity to investigate the relationship between mosquito dynamics and dengue outbreaks using mathematical models. With this aim, in this paper, we propose an enhanced deterministic model assuming functions of temperature and time for the entomological parameters to analyze and simulate the temporal dynamic of dengue in Foz do Iguaçu, evaluating the influence of climate on the vector population and their consequent impact on the dengue incidence.

## 2 Materials and Methods

### 2.1 Data description and processing

Foz do Iguaçu is situated in the Western part of the Paraná State, Brazil, at 25°32’49 S latitude and 54°35’18” W longitude. The city stands at an elevation of 164 meters above sea level and covers a total area of 618.352 km^2^. The mean annual temperature in the region is 20.4°C, and the annual average rainfall amounts to 1,800 mm. The municipality is home to a population of 256 thousand inhabitants and has a HDI of 0.751 [20].

#### 2.1.1 Epidemiological data

The linelist of anonymized dengue cases by local of residence in Foz do Iguaçu was obtained from Notifiable Diseases Information System (SINAN), [5], provided by Foz do Iguaçu Health Department. The dataset contains daily georeferenced information on reported cases, for the period between 07/01/2010 and 30/12/2022. In this period, 23.52% of the notified cases were lab-confirmed, and 2.05% had serotype information.

#### 2.1.2 Entomological data

Data on mosquito traps are aggregated according to high-quality bimonthly collection. We use data (odd months) from 09/01/2017 to 13/05/2022, totaling 33 collections about trapped mosquitoes provided by the city’s Zoonosis Control Center (CCZ). There are 2500 georeferenced mosquito traps (Adultrap) distributed throughout the city. The trap is designed to attract and capture female mosquitoes seeking oviposition sites. The entomological data came from the Foz do Iguaçu citywide vector surveillance program that bimonthly deploys adult mosquito traps throughout the city.

We calculated the MFAI (Mean Female *Aedes sp*. index) as the ratio between the number of females captured and the number of traps inspected [25]. Based on observations from Foz do Iguaçu field entomologists, mosquito counts per trap are considered reliable only for those captured within a maximum of 15 days prior to the collection date. Mosquitoes collected earlier than this period are at a significant risk of deterioration or predation. Consequently, for model fitting purposes, we assume that the MFAI metric represents the mosquitoes captured in the trap within the last 15 days.

#### 2.1.3 Climate data

Meteorological data collected by the Meteorological Station at Foz do Iguaçu was obtained from Sistema Meteorológico do Paraná (SIMEPAR) (http://www.simepar.org/prognozweb/simepar). It contains daily information about the average and the minimum and maximum temperatures, precipitation, average humidity and wind speed for the period between 01/01/2010 and 30/12/2022. Meteorological data with missing values were filled using the average of the previous seven days.

### 2.2 Mathematical modelling

#### 2.2.1 Complete dengue model

Aiming to mathematically describe dengue transmission dynamics in Foz do Iguaçu, we departed from the deterministic model proposed by [35], referred here as *complete dengue model* (Fig. 1, eq. 1). The model describes the transmission of a single dengue serotype between mosquitoes and humans in a homogeneous setting where *M* consists of the adult female mosquito population and *A* aquatic stage mosquitoes. The total number of adult females, *M*, is divided into three subdivisions: susceptible *M*_*s*_, exposed *M*_*e*_ and infected mosquitoes *M*_*i*_.

**Figure 1:**
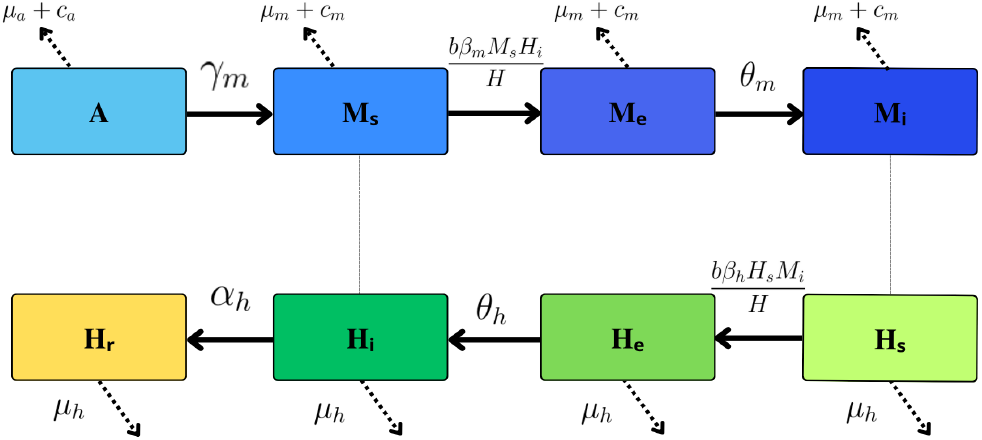
Diagram of the complete model proposed by [35] showing the population compartments and the transition rates.

As for the total human population, it is considered constant and has four compartments: susceptible *H*_*s*_, exposed *H*_*e*_, infectious *H*_*i*_ and recovered *H*_*r*_. Thus, we have *M* = *M*_*s*_ + *M*_*e*_ + *M*_*i*_ and *H* = *H*_*s*_ + *H*_*e*_ + *H*_*i*_ + *H*_*r*_.

Figure 1 shows a diagram of the complete dengue model. The entomological parameters that assume different values for different temperatures are: *δ*(*T* (*t*)) is the oviposition rate, *γ*_*m*_(*T* (*t*)) is the rate at which mosquitoes emerge from the aquatic phase, *µ*_*a*_(*T* (*t*)) and *µ*_*m*_(*T* (*t*)) are the aquatic and adult phase mosquito mortality rate, respectively. *C*(*t*) is the mosquito carrying capacity, assumed constant, but later this assumption was relaxed, as seen in the results. The *c*_*a*_ and *c*_*m*_ are death rates of immature and adult mosquitoes, respectively, induced by control efforts. The human mortality rate is written as *µ*_*h*_. We denote the average number of bites per mosquito per day as *b*, the per capita mosquito biting rate. We consider *β*_*m*_ and *β*_*h*_ as the transmission probabilities from human to mosquito and from mosquito to human, respectively. The parameters *θ*_*m*_ and *θ*_*h*_ are the rates at which mosquitoes and humans become infectious and their reciprocal quantities, 1*/θ*_*m*_ and 1*/θ*_*h*_ are known as extrinsic and intrinsic incubation periods, respectively.

We reproduce (1), the system of non-linear differential equations of the complete model [35].

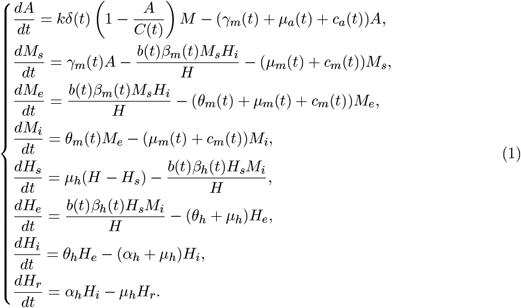

To fit this model to Foz do Iguaçu’s data, it was necessary to modify some of its assumptions, as explained in subsection 2.2.3.

#### 2.2.2 Mosquito capture model

We proposed a sub-model containing only the mosquito population, without infection to fit the mosquito component of the complete model with available trap data, refereed as *mosquito capture model*. In this model, we additionally represented the trapping process, in a similar way to [24]. The population of trapped mosquitoes is included in the model as an extra equation, however the variables and parameters are the same as in the *complete model*.

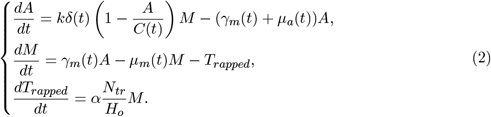

Note that the *mosquito capture model* is a particular case of the *complete model* for which there is no infection; due to that, there are only the aquatic and adult phases of the mosquito population. The third equation counts the captured mosquitoes, where the variable *T*_*rapped*_(*t*) represents the accumulated number of mosquitoes captured during the surveillance period from time 0 until t.

To fit the model with data from mosquitoes captured in each bimonthly collection, we calculate the MFAI as in [24], which is the ratio between the number of females captured and the number of traps inspected in the collection. The theoretical MFAI is calculated every two months *t*_*b*_ by the equation,

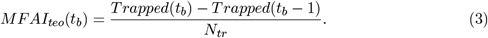

#### 2.2.3 Temperature-dependent entomological parameters

Following [44], we assume that the entomological parameters show temporal variation associated with the daily temperature in Foz do Iguaçu. Table 1 describes the complete model parameters including the entomological ones, as well as the range of values.

**Table 1:**
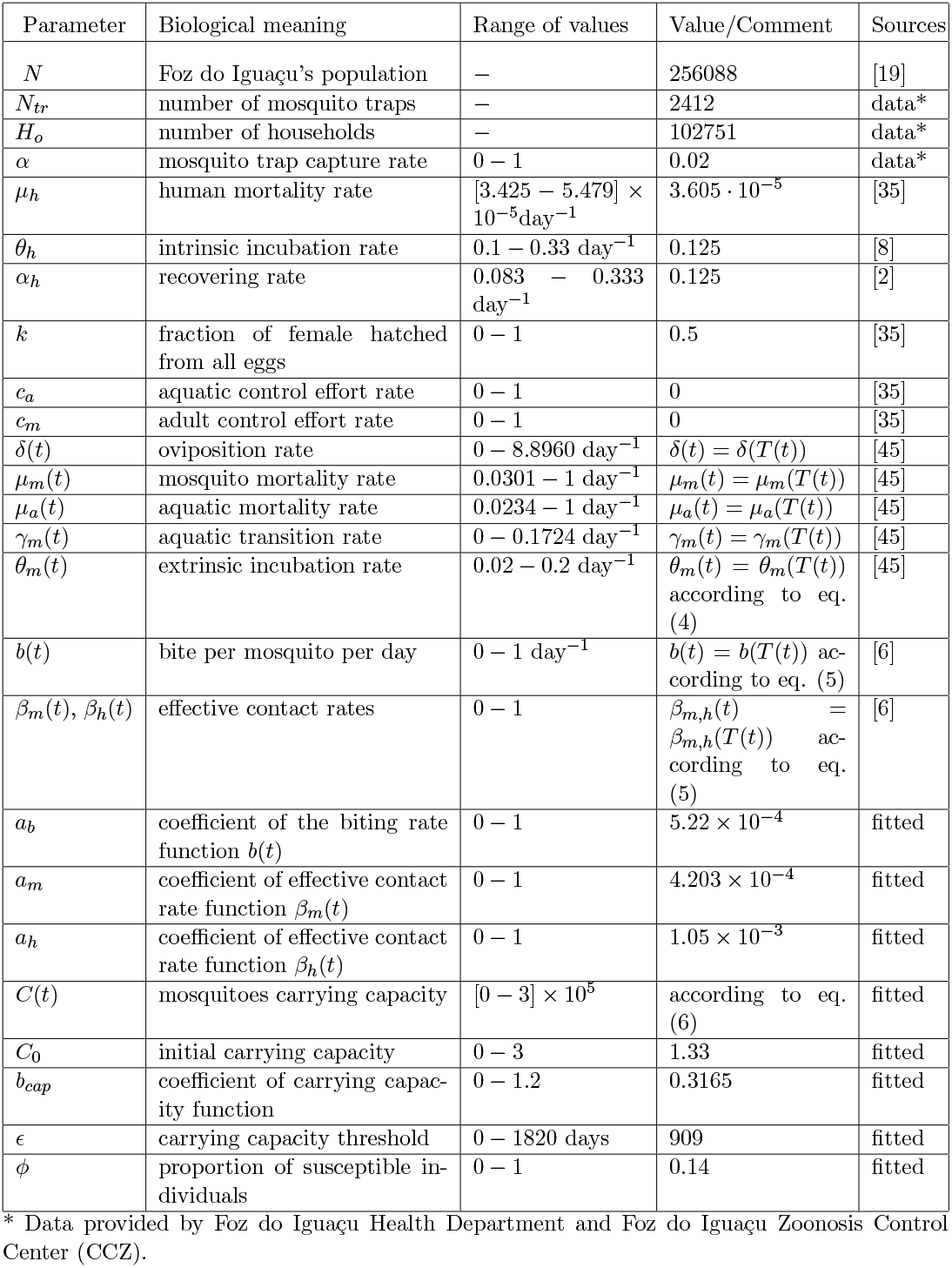
Complete dengue model: symbols, description, values, and references.

We used the values for five entomological parameters that vary with temperature, *δ*(*t*), *µ*_*m*_(*t*), *µ*_*a*_(*t*), *γ*_*m*_(*t*) and *θ*_*m*_(*t*) proposed by [45]. The expressions for parameters *δ, µ*_*m*_, *µ*_*a*_, *γ*_*m*_ are obtained in [45] and [44], to a polynomial of degree n, *P*_*n*_(*T*) = *b*_0_ + *b*_1_*T* + · · · + *b*_*n*_*T* ^*n*^. Now, for the extrinsic incubation *θ*_*m*_, according to [27], it depends on the temperature as follows:

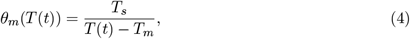

where *T* (*t*) is the average daily temperature in degrees Celsius °C, *T*_*s*_ is the thermal sum (°C × day^−1^), measured in degree-days representing the accumulation of temperature units over time, and *T*_*m*_ is the threshold of temperature below which dengue virus cannot multiply, hence *T > T*_*m*_. We assume *T*_*m*_ = 14°*C* and *T*_*s*_ = 135°*C × day*^−1^ according to [13].

Using the above expressions, and the daily temperature of Foz do Iguaçu as input, we calculated the daily varying entomological parameters for the model.

Between 2010 and 2022, the minimum temperature in Foz do Iguaçu was *T*_*min*_ = −1.8°*C* and the maximum *T*_*max*_ = 41.8°*C*. Parameters extrapolations were proceeded based on biological literature regarding mosquito survival outside these ranges from [44]. Different temperature intervals are defined for the interpolation of each parameter. Except for *θ*_*m*_, which, due to its formulation, does not allow values of *T* ≤ 14°*C*. Parameter values for temperatures outside the interpolation range were fixed at their extreme values within the range as follows.

It is important to address the borders from the entomological parameters ranges of variation with temperature. For the oviposition rate, from [44], there is not any oviposition for the following temperatures: *δ*(*T* ≤ 10.8°*C*) = *δ*(*T* ≥ 37.3°*C*) = 0.

Concerning the aquatic transition rate *γ*_*m*_, for the following temperature ranges, there is not aquatic transition because the aquatic mortality rate is high and the oviposition rate is almost zero so the adjusted polynomial crosses the temperature axis on: *γ*_*m*_(*T* ≤ 12.0°*C*) = *γ*_*m*_(*T* ≥ 40.3°*C*) = 0. For the average aquatic mortality rate *µ*_*a*_, from data on [44], larva mortality is very high for low and high temperatures, so: *µ*_*a*_(*T* ≤ 5.2°*C*) = 1.0. For the average adult mortality rate *µ*_*m*_, we see from the data on [44] that for very low temperatures, the mortality *µ*_*m*_(*T* ≤ −0.9°*C*) = 1.0. For *b, β*_*m*_, *β*_*h*_ is *b, β*_*m*_, *β*_*h*_(*T* ≤ 18.0°*C*) = *b, β*_*m*_, *β*_*h*_(*T* ≥ 34.0°*C*) = 0. The extrinsic incubation rate *θ*_*m*_ is defined as *θ*_*m*_(*T* ≤ 16.4°*C*) = 0.02 e *θ*_*m*_(*T* ≥ 38°*C*) = 0.2.

The temperature dependence of the parameters *b*(*t*), *β*_*m*_(*t*), and *β*_*h*_(*t*) is described by the asymmetric Brière function [30]:

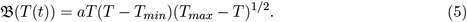

Following [30], we set for *b*(*t*) the following temperatures: *T*_*min*_ = 13.35°*C* and *T*_*max*_ = 40.08°*C*. Now for *β*_*m*_(*t*) we have *T*_*min*_ = 12.22°*C* and *T*_*max*_ = 37.46°*C* and for *β*_*h*_(*t*), *T*_*min*_ = 17.05°*C* and *T*_*max*_ = 35.83°*C*. Assuming that virus transmission occurs only within this temperature range. In cases where 𝔅(*T* (*t*)) *<* 0, this value is replaced by zero. The coefficient *a* is fitted in the complete model for the three parameters *b*(*t*), *β*_*m*_(*t*), and *β*_*h*_(*t*) (see section 2.3). Figure 2 shows the behavior of the three parameters described by the Brière function.

**Figure 2:**
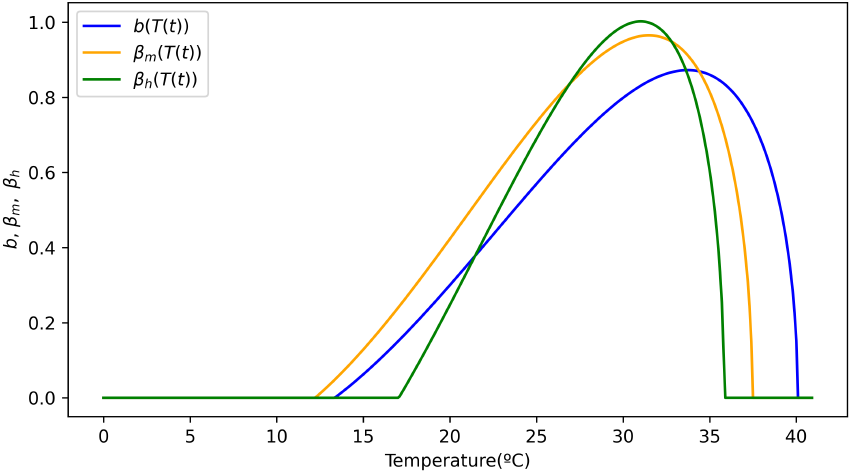
Parameters behavior *b*(*t*), *β*_*m*_(*t*), and *β*_*h*_(*t*) with temperature according to the Brière functions with fitted parameters. *a*_*b*_ = 5.0366 · 10^−4^, *a*_*m*_ = 6.5093 · 10^−4^, *a*_*h*_ = 1.0546 · 10^−3^.

#### 2.2.4 Carrying capacity

Thus, for the carrying capacity parameter, it is observed that there was a significant increase in captured mosquitoes from 2019 onwards during the period 2017-2021 (figure 3). It seems there is no significant temperature or precipitation variation that justifies this increase, and furthermore, the number of traps analyzed throughout the period was nearly constant in the study period.

**Figure 3:**
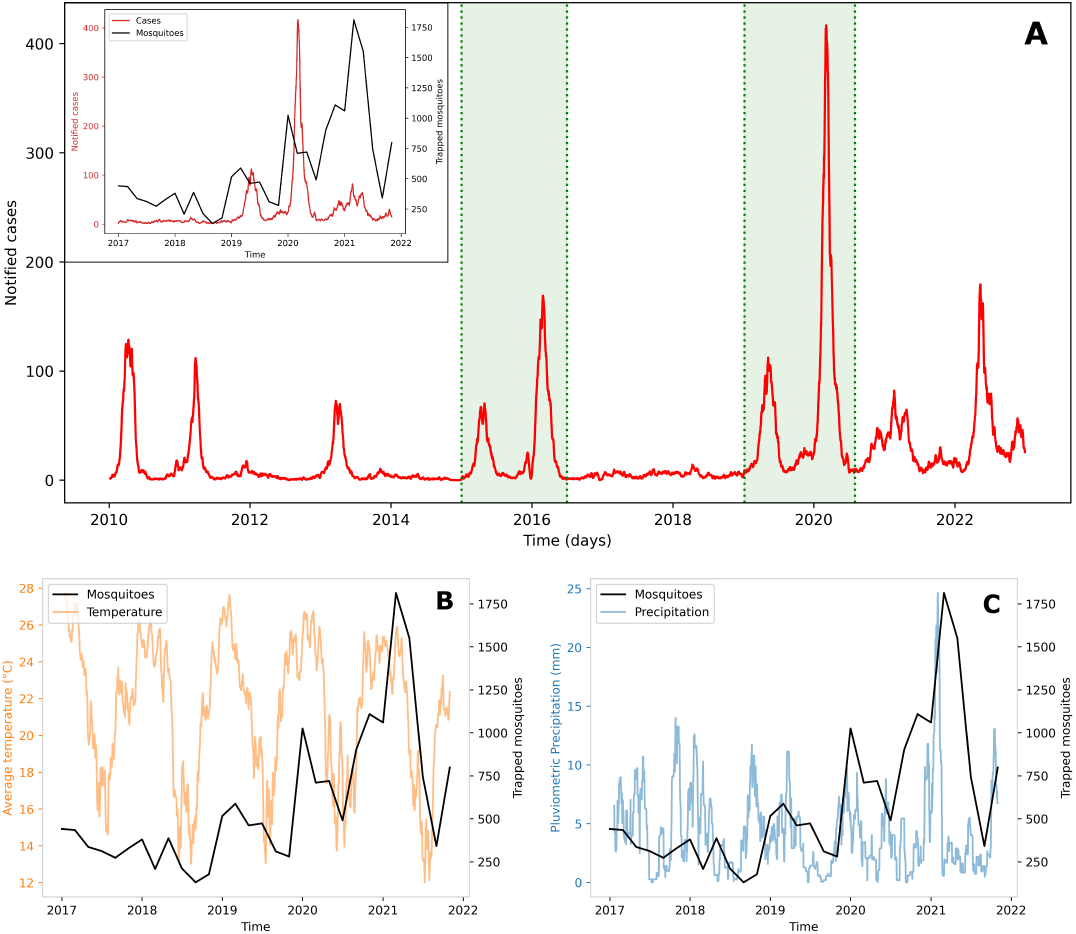
Panel showing the datasets used in this study: a) Time series of dengue cases in Foz do Iguacu Brazil from 2010 to 2022. The shaded green area represents the periods used for curve fitting. The inset graph shows both trapped mosquito and dengue cases, for comparison from 2017 to 2022. b) Time series of bimonthly trapped mosquitoes and temperature, with a two-week moving average, for the period we have mosquito data. c) Time series of bimonthly trapped mosquitoes and precipitation, with a three-week moving average, for the period we have mosquito data.

**Figure 4:**
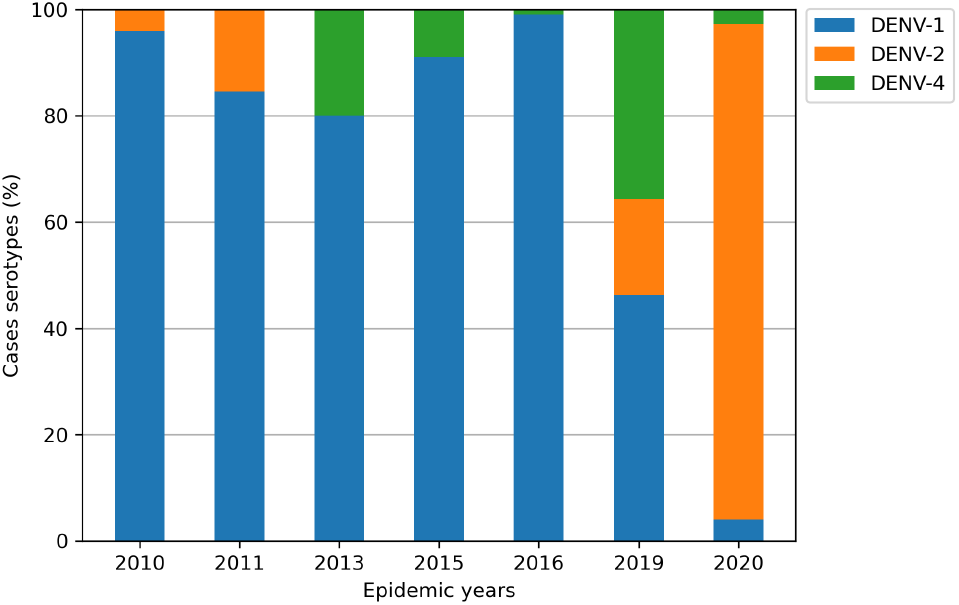
Description of the predominant DENV serotypes between 2010 and 2020 in Foz do Iguaçu, Brazil. The figure shows DENV-1 was prevalent between 2010 and 2016 while we observed a strong detection of DENV-2 in 2020.

To achieve the formulation of the carrying capacity, different models were tested, as proposed by [44], considering ranges of temperature and precipitation. Also, we consider a linear relationship with precipitation according to [36]. The best formulation was the one which provided the best goodness-of-fit with the captured mosquito data following a temporal dependence without explicit dependence with climate variables, the Heaviside function.

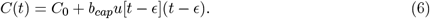

In the expression (6), *C*_0_ represents the initial carrying capacity, *u*(*t*) is the Heaviside function, *b*_*cap*_ is a constant rate associated with the growth of carrying capacity, and *ϵ* is a constant parameter that represents the time at which the carrying capacity begins to increase. *C*(*t*) assumes values of the order of 10^4^.

Based on the above considerations about the entomological parameters and on the literature about other parameters, we complete table 1 with the values range of the *complete model*, including the ones presented by the temperature-dependent functions of entomological parameters.

#### 2.2.5 Mosquito capture model parameters

For the mosquito model, we assume the trapping process described by *αN*_*tr*_*/H*_*o*_, where *α* is the trap attractiveness (attracted mosquito proportion by the trap within a household), and *N*_*tr*_ and *H*_*o*_ are the number of traps and households in Foz do Iguaçu, respectively. Therefore, the ratio *N*_*tr*_*/H*_*o*_ is the trap density per household. We assume *α* = 0.02 arbitrarily, meaning that 2% of mosquitoes in a trap are captured per day. This value is chosen as previous studies [39] indicate a low capture rate.

### 2.3 Sensitivity analysis and model calibration

A sensitivity analysis from the Sobol method ([15]) was conducted to decide which parameters should be fitted. In the mosquito trapped model, we focused on parameters *δ, γ*_*m*_, *µ*_*a*_,*µ*_*m*_, *C*_0_, *ϵ* and *b*_*cap*_ to analyze their impact on the total number of mosquitoes trapped (2). In the complete dengue model (1), we examined how the model is sensible to the parameters *a*_*b*_, 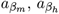, *C*_0_, *α*_*h*_, and *θ*_*h*_ regarding the total number of infected humans.

After analyzing the sensitivity, the parameters *C*_0_, *b*_*cap*_, *ϵ, ϕ a*_*b*_, 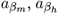,*a*_*h*_, were fitted in the models. To achieve this, we use the *lmfit* package of Python, based on the *Levenberg-Marquardt* (damped least-squares) fitting method [32].

For the mosquito model calibration, we use the trap data to estimate the coefficients *C*_0_, *b*_*cap*_ and *ϵ* of the carrying capacity equation (eq. (6)).

With the fitted mosquito model, we proceed to fit the parameters *ϕ, a*_*b*_, *a*_*m*_, *a*_*h*_, *C*_0_, of the complete model, using the time series of cases, where *ϕ* refers to the proportion of susceptible individuals and *a*_*b*_, *a*_*m*_, *a*_*h*_ correspond to the parameter *a* in the respective Brière functions (5) for *b*(*t*), *β*_*m*_(*t*), and *β*_*h*_(*t*) and *C*_0_ the initial carrying capacity.

When we fit the model to the data, the following expression is minimized:

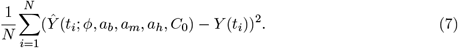

Here, *Ŷ*(*t*_*i*_) represents the new infections (*θ*_*h*_*H*_*e*_(*t*_*i*_)) from the complete model, and *Y* (*t*_*i*_) represents the daily reported new cases data in the same period.

### 2.4 Reproduction number of dengue in Foz do Iguaçu

The basic reproduction number *R*_0_ is an important indicator of transmission, that measures how fast the number of cases increases, with values larger than 1 implying when there is an epidemic process. Here, we use the expression for the dengue basic reproduction number, *R*_0_, as derived from the complete model [35] using the next-generation matrix method. This calculation assumes an exponential increase in cases at the beginning of an epidemic. The expression for *R*0 is presented in equation (8):

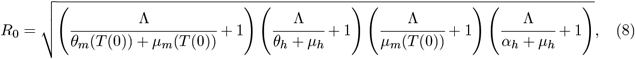

where the force of infection Λ corresponds to the exponent of new cases at the beginning of the epidemics.

To calculate *R*_0_ from data, we need to define the time interval for which there is exponential growth. For that time interval, it results in a linear growth in the plot of new cases against the cumulative number of cases.

After the initial growth, the temporal evolution of the epidemic is not constant and its velocity is measured by the time-dependent reproduction number, *R*(*t*). An expression for *R*(*t*) was derived from the complete model by [35] using the method described in [43]. Thus, *R*(*t*) is estimated by:

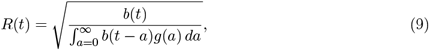

where *b*(*t*) is the number of new cases at week *t* and *g*(*a*) is the dengue generation time interval distribution, which is defined as the probability distribution of the time an infected individual takes to infect a secondary case. In [35], following [3], the expression is

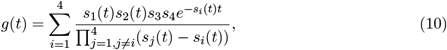

where *s*_1_(*t*) = *θ*_*m*_(*t*) + *µ*_*m*_(*t*) + *c*_*m*_(*t*), *s*_2_(*t*) = *µ*_*m*_(*t*) + *c*_*m*_(*t*), *s*_3_ = *θ*_*h*_ + *µ*_*h*_, *s*_4_ = *α*_*h*_ + *µ*_*h*_. The parameters and their values are presented in table 1.

Unlike [35], which utilized fixed temperature values for the entomological parameters, our analysis for Foz do Iguaçu considers the parameters *θ*_*m*_(*t*) and *µ*_*m*_(*t*) as functions of the observed temperature data.

## 3 Results

The results will be presented in four subsections. First, we show time series data for dengue cases and trapped mosquitoes with temperature and precipitation. Then, we present the mosquito dynamics model in section 3.2, followed by the dengue dynamics in section 3.3 and, at last, the reproduction number in section 3.4.

### 3.1 Dengue cases and trapped mosquitoes

The time series data from mosquitoes and dengue cases in Foz do Iguaçu are illustrated in figure 3. Figures 3b and c specifically show the time series for trapped mosquitoes, along with temperature and precipitation, respectively.

From 2010 to 2021, Foz do Iguaçu experienced nine dengue outbreaks, in the years 2010, 2011, 2013, 2015, 2016, 2019, 2020, and 2021. For modeling, we selected the 2015-2016 and 2019-2020 outbreaks (highlighted by green shaded areas in figure 3) because these epidemic years exhibited double peaks. This pattern is likely due to the predominant presence of a single serotype, which aligns with one of the assumptions in our model. Entomological data are available only for the 2019-2020 epidemic period.

Furthermore, the occurrence of different DENV serotypes over the years is shown in 4.

### 3.2 Mosquito dynamics

First, we performed a local sensitivity analysis where each parameter was varied while all the others were fixed. Then, a Sobol analysis was performed and the output is in figure 5.

**Figure 5:**
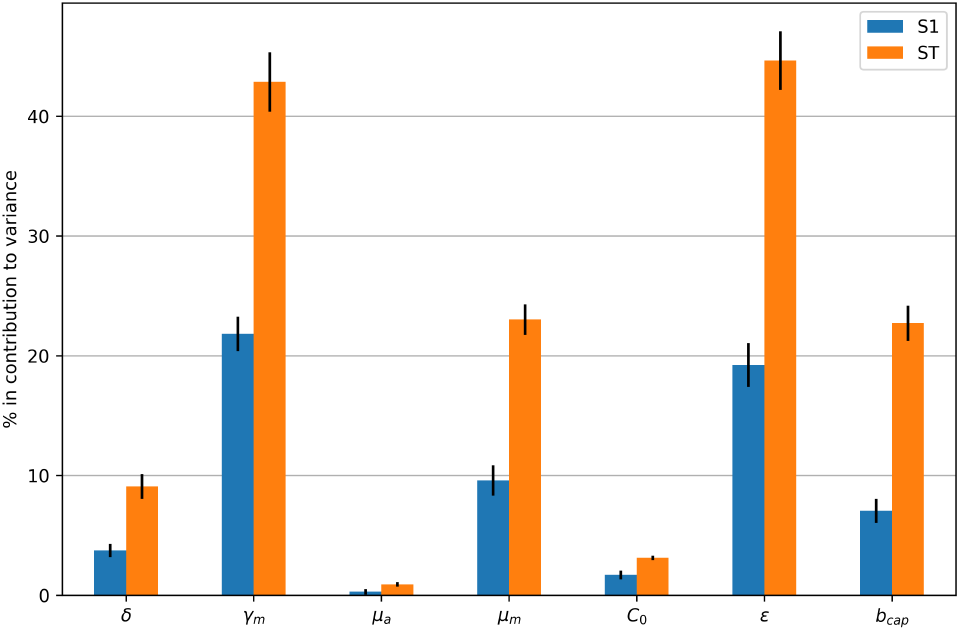
Sensitivity of mosquito total number of trapped mosquitoes to model parameters. S1 and ST represent first-order and total sensitivity indices, respectively. The parameters and scanned intervals for this analysis were: *δ*: [0,9]day^−1^, *γ*_*m*_: [0, 0.2]day^−1^, *µ*_*a*_: [0.0234,0.5]day^−1^, *µ*_*m*_: [0.0301, 0.109]day^−1^, *C*_0_: [10, 100], *ϵ*: [0, 1000], *b*_*cap*_: [0.001, 1.2].

The parameters that affects the number of trapped mosquitoes the most were the carrying capacity threshold (*ϵ*) and the aquatic transition rate (*γ*_*m*_).

#### 3.2.1 Fitting and numerical simulations

For the initial conditions of the mosquito model, we assumed *M* (0) = 0.7*N* where *N* = 256, 088 is the total population of Foz do Iguaçu. A similar mosquito-to-human ratio was the same used by [4], based on measures from [31]. As for the aquatic phase, it was assumed *A*(0) = 0.85*C*_0_, with *C*_0_ being the carrying capacity at *t* = 0, as estimated by the model. The initial number of trapped mosquitoes was considered *T*_*rapped*_(0) = 0.

In order to fit the parameters *C*_0_, *b*_*cap*_ and *ϵ* of the carrying capacity *C*(*t*)(*eq*.6) we use the captured mosquito data. The model fits the data well, predicting its seasonality and growth trend (figure 6).

**Figure 6:**
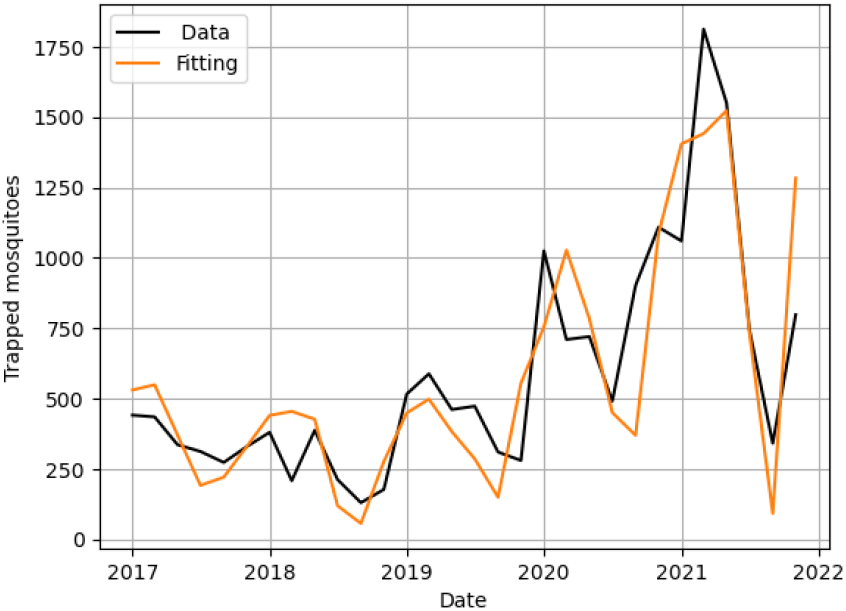
Observed data and simulated curve of mosquito model with the following fitted parameter values: *C*_0_ = 1.27 · 10^5^, *b*_*cap*_ = 0.3165, *ϵ* = 909 days.

### 3.3 Dengue dynamics

Here we consider the complete dengue model (1) based on the simulated results of the mosquito model, performing a sensitivity analysis and fitting the model with notified dengue cases.

Figures 5 and 7 display the contributions of these parameters to the model, where S1 and ST represent the first-order, and total sensitivity indices, respectively. These indices provide a decomposition of output variance relative to each parameter. Notably, if the total-order indices (ST) are significantly larger than the first-order indices (S1), it indicates the presence of higher- order interactions among the parameters.

**Figure 7:**
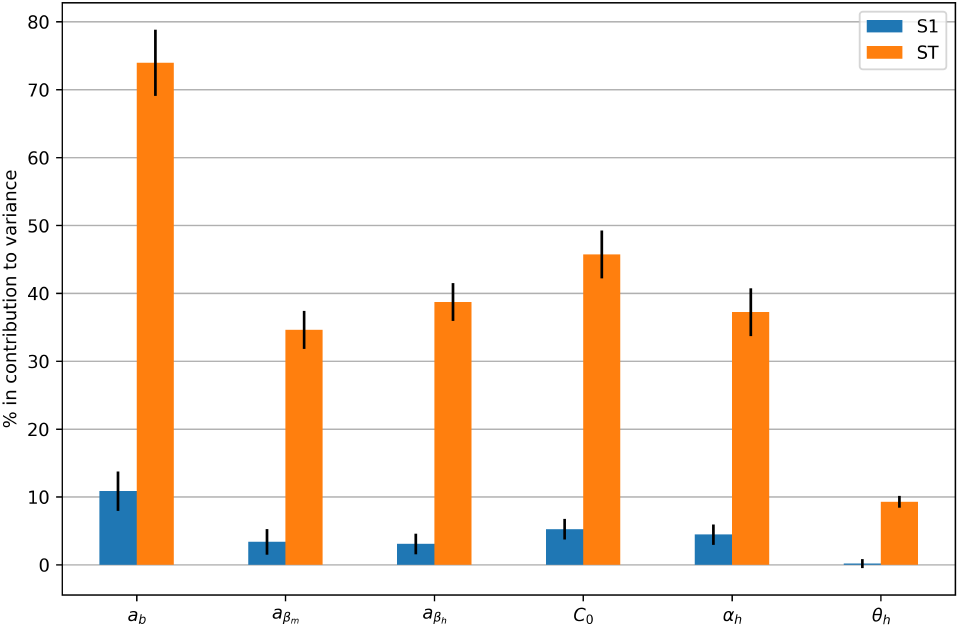
Sensitivity analysis of complete model parameters with respect to total infected humans. S1 and ST represent first-order and total sensitivity indices, respectively. The parameters and scanned intervals for this analysis were: *a*_*b*_ : [0.0007, 0.00138], 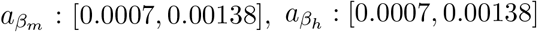, *C*_0_ : [12, 28], *α*_*h*_ : [0.083, 0.25]*day*^−1^, *θ*_*h*_ : [1*/*12, 1*/*3]*day*^−1^.

When integrating the complete model, the total number of simulated infected humans is used as input to assess the model’s sensitivity to each parameter. The results of this sensitivity analysis are shown in figure 7. *a*_*b*_ associated with the bite rate *b* is the most significant parameter with respect to the total number of infected humans.

### 3.3.1 Fitting and numerical simulations

To fit the model to the dengue data during the 2015-2016 outbreaks, we used data from 01/01/2015 to 01/07/2016. The parameters *ϕ, a*_*b*_, *a*_*m*_, *a*_*h*_ and *C*_0_ were fitted. During both seasons considered here, we did not have mosquito population data to estimate C(t) so we assume that the carrying capacity was constant following the expression *C* = *C*_0_·0.7*N*. The initial conditions were: *A*(0) = 0.85·*C*(0) = 1.85×10^5^, *M*_*s*_(0) = 0.7·*N, M*_*e*_(0) = *M*_*i*_(0) = 0.7·*H*_*i*_(0) = 13, *H*_*s*_(0) = *N* − *H*_*e*_(0) − *H*_*i*_(0) − *H*_*r*_(0) = 19243, *H*_*e*_(0) = 18 = *H*_*i*_(0), *H*_*r*_(0) = (1 − *ϕ*)*N* = 236881.

As for the 2019-2020 seasons, we used data from 06/01/2019 to 30/12/2020. The model was fitted calibrating *ϕ, a*_*b*_, *a*_*m*_, *a*_*h*_, *C*_0_. For these outbreaks, the initial conditions for the mosquito compartment are obtained from the fitted mosquito model: *A*(0) = 1.33 × 10^5^, *M*_*s*_(0) = 3.63 × 10^5^. For the human compartment, we set *M*_*e*_(0) = *M*_*i*_(0) = 0.7*H*_*i*_(0) = 43, *H*_*s*_(0) = *N* −*H*_*e*_(0) − *H*_*i*_(0) − *H*_*r*_(0) = 31433, *H*_*e*_(0) = *H*_*i*_(0) = 62, *H*_*r*_(0) = (1 − *ϕ*)*N* = 226979. In table 1, the range of values of the model parameters and the goodness-of-fit parameters estimated in the complete model for 2019-2020 are presented.

The dengue model captured well the seasonality of dengue in both periods. There was an overshoot in the 2016 outbreak, but all the other three were correctly described by the model.

### 3.4 Basic and Effective r eproduction numbers f or e pidemic years

The estimated values of *R*_0_ and Λ for the epidemic years in Foz do Iguaçu are shown in table 2. The precision of the *R*_0_ estimation is better during epidemic years where there is a well-defined exponential growth. The largest estimated values were found for the years 2010 and 2020. Figure 9 shows the linear phase for these years.

**Table 2:**
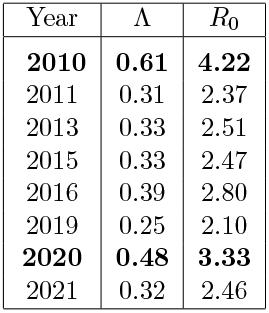
Force of infection and basic reproduction number by epidemic year in Foz do Iguaçu, Brazi.

In figure 9, we show the force of infection and basic reproduction number for the 2010 and 2020 epidemic years in Foz do Iguaçu. The two periods highlighted in bold coincide with the entrance of serotype DENV-1 and DENV-2 respectively, where we can see the initial exponential growth as fitted by a straight line, using data and equation. (8).

To calculate the time-dependent reproduction number *R*(*t*), we consider the following dependence with temperature for parameters *θ*_*m*_, *µ*_*m*_ which takes part into the generation interval *g*(*t*) composing the rates of leaving the exposed and infectious compartments *s*_1_(*t*) and *s*_2_(*t*). The result of the calculated effective reproduction number can be seen in figure 10, where we observe the instantaneous effect of temperature, in comparison to its value for an average temperature. The results suggest that high heat waves may promote large time intervals for which *R*(*t*) *>* 1. Also, note its high sensitivity to small variations of the weekly new cases and a tendency to grow near the outbreaks, as expected of a reproduction number, presenting the maximum *R*(*t*) values associated with 2020 and 2022 outbreaks.

## 4 Discussion

In this work, we analyze the expansion of dengue in Foz do Iguaçu through a mathematical modeling perspective, counting a rich dataset of mosquito and human infection data. This city has experienced a sequence of dengue outbreaks in the last two decades, as seen in figures 3a and 3b. In response, Foz do Iguaçu has implemented a comprehensive entomological surveillance program since 2017 [14], integrated with a sensible and responsive disease surveillance program. During the study period, the largest epidemics occurred in 2016 and 2020, both preceded by a shorter epidemic in the previous year. Sequences of epidemics like this suggest that transmission was interrupted during the winter to return in the next year. A pattern of two subsequent waves has been observed in other cities, like Taiwan [9], and Rio de Janeiro [34], and has been attributed to multiple serotypes or viral evolution.

Our analyses indicate that a one-serotype model with temperature as external forcing can produce the two-wave pattern observed in the data, with the second peak larger than the first. That is, the effect of temperature on mosquito dynamics in Foz do Iguaçu is sufficient to induce the observed dengue seasonality, even in the absence of a new serotype.

The sensitivity analyses of both, the mosquito and complete models reveal that climate- dependent parameters, such as carrying capacity *C*(*T*), aquatic transition rate *γ*_*m*_ and bite rate *b*(*T* (*t*)), are the most sensitive in relation to mosquitoes’ population and dengue incidence. Since the bite rate is multiplied by the infection rates between humans and mosquitoes, its action, combined with the infection rates, affects the nonlinear terms of the complete model responsible for the interaction between humans and mosquitoes, considering both approaches related to the vector population and the human infection load.

Another important result from this study refers to the mosquito dynamics. The data suggests that mosquito abundance increased sharply from 2020 onwards. Initial versions of the model with constant or seasonal carrying capacity, were not capable of representing this increasing abundance. Only a model with a linear trend of the carrying capacity correctly captured the observed pattern. Increasing carrying capacity indicates a relaxation of the density-dependence effects on mosquito population growth. Such a fact can occur if the number of breeding sites increases, thus reducing the competitive pressure in the aquatic stage. It is noteworthy that the 2020 - 2022 period corresponds to the first two years of the COVID-19 pandemic, during which significant changes occurred in human behavior and vector control efforts. Similar observations have been made in other locations [38]. We hypothesize that these changes may have contributed to alterations in mosquito abundance in Foz do Iguaçu.

We also investigated the temporal dynamics of dengue transmission and its relationship with climate variables, such as temperature, considering that it reflects much more than seasonal features. Through the complete model (1), [35], we assumed the dependence on the average temperature in the entomological parameters vary with daily temperature according to some considered temperature-dependent functions [44, 45].

Through the model, we could confront data about human infection generated by simulation and reported dengue cases from surveillance data (see figure 8b),considering the mosquito population and climate effect, as previously demonstrated by [26]. The numerical results confidence of the dengue complete model [35] is based on the confrontation of the mosquito population dynamics model with observed data (see figure 6) of the trap model to set up realist entomological parameter values to be used in the complete model.

**Figure 8:**
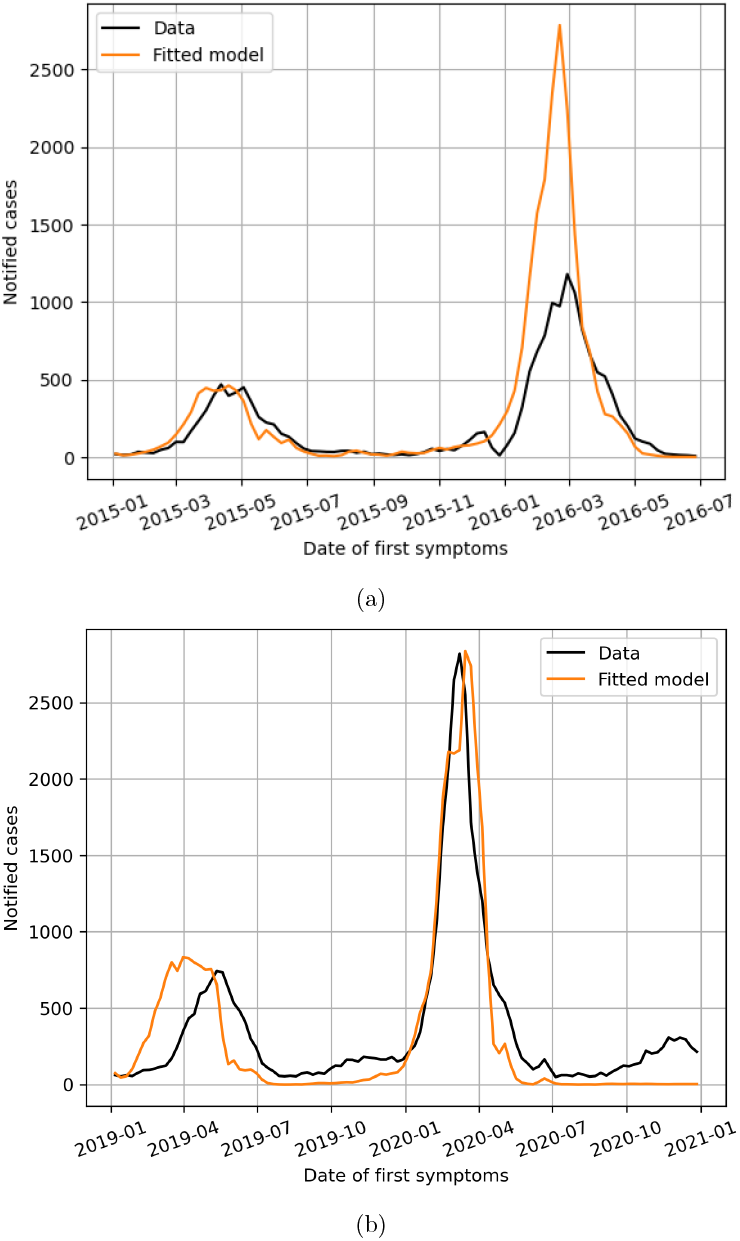
Fitting complete model to data. The black line represents the data of new weekly cases, and the orange line represents the curve estimated by the model. The goodness-of-fit model parameters for the outbreak of 2015-2016 (a) are: *ϕ* = 0.075, *a*_*b*_ = 5.316 × 10^−4^, *a*_*m*_ = 6.616 × 10^−4^, *a*_*h*_ = 1.06 × 10^−3^, *C*_0_ = 0.999 with constant carrying capacity. For the outbreak of 2019-2020 (b), we have: *ϕ* = 0.14, *a*_*b*_ = 5.22 × 10^−4^, *a*_*m*_ = 4.203 × 10^−4^, *a*_*h*_ = 1.05 × 10^−3^, *C*_0_ = 1.33 presented in Table 1.

**Figure 9:**
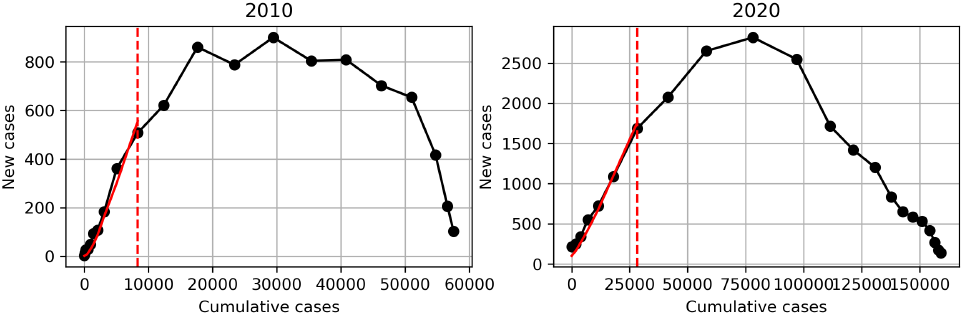
The black line represents the data of new cases against the cumulative number of cases, the red dashed line represents the end of the linear phase, and the red solid line represents the estimated curve. On the left side (a), it is shown data for 2010 dengue epidemics, and on the right side (b), data for 2020.

**Figure 10:**
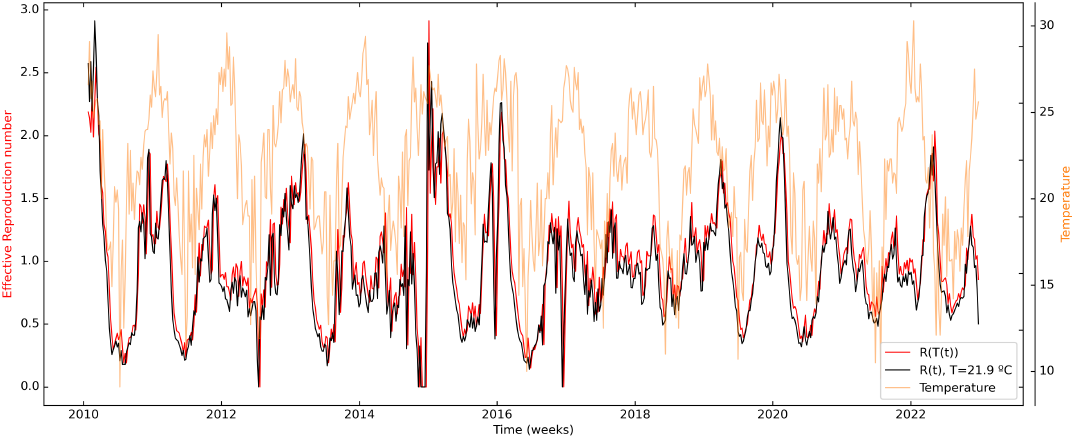
Effective reproduction numbers (eq. 9) with and without temperature dependence. Each parameter value used is present in table 1 and is multiplied by seven for weekly values.

The basic reproduction number, *R*_0_, shows higher transmission in 2010 compared to 2020. This result reinforces neither the number of cases nor the incidence captures the information revealed in *R*_0_, which is associated with the impact of transmission during the beginning of the epidemic. Some hypotheses can explain this phenomenon. First, this observation occurs in years of new viral variants introduction, DENV-1 in 2010 and DENV-2 in 2020, as shown by data from dengue serological surveillance. Another hypothesis is that *R*_0_ estimations are more associated with the ecological characteristics of the vector associated with the circulation of non-specific serotypes ([33]). Another hypothesis, as discussed by [18], is the association between the timing of the start of interventions and the epidemic magnitude.

In this study, we also compared the estimation of *R*(*t*) using expressions that either include or do not include the temperature data. Despite the small difference in magnitude between the two estimates, our results indicate the expression incorporating temperature consistently produces higher values of *R*(*t*) compared to the one that does not. This implies the temperature-inclusive expression will more frequently exceed the transmission threshold (*R*(*t*) *>* 1), making it a more sensitive measure of *R*(*t*) for use in alert systems. These findings are consistent with the observations reported by [10].

As the disease progresses, several factors, such as control measures and changes in the susceptible population can alter the scenario of disease dynamics so that it is less reasonable to consider its development in a constant environment with exponential growth of cases. Therefore, the time-dependent effective reproduction number, *R*(*t*), captures the temporal evolution of the reproduction number. Furthermore, ecological factors, urbanization, population mobility, deforestation and insecticide resistance, together with climatic events and anomalies in recent decades, have created conditions for pathogens and arbovirus vectors to emerge in new areas or re-emerge in regions, even with powerful antimicrobial campaigns. [42], [7], [1], [17]. These heterogeneous characteristics also lead to different epidemic scenarios according to susceptibility and other factors [21].

## 5 Final considerations

In conclusion, our results call attention to the relevance of mosquito trap data in constructing a confidence time series of mosquito population dynamics to get a more precise scenario of dengue dynamics. Both fitted models for Foz do Iguaçu display good fitting to data, showing a promising methodology of performing a previous fitting of the entomological parameters in a population mosquito model and then applying it to the dengue model. Particularly, this analysis may provide elements to help the description of dengue dynamics for other regions with subtropical climates where dengue infection is expanding. They can be used to plan interventions on pre-epidemic moments and generate timely alerts.

The results also highlight the role of climate variables, particularly temperature, directly affecting mosquitoes’ entomological characteristics or modifying human behavior, which, in turn, may impact the mosquito population. However, many other factors are relevant for the understanding of the complex dengue; in that direction, our results also emphasize that the climate alone does not explain either the increase of mosquitoes or the expansion of dengue.

Therefore, studies taking into account the link between modeling and data as well as the inclusion of other elements in the model. Co-circulation of serotypes [11] and other diseases also transmitted by the same vector (Zika and chinkugunya) [16] may help to lead to a more complete description of dengue dynamics and, as therefore, a more efficient design of control scenarios with vaccine and vector control procedures.

## Data Availability

We confirm that all database and code used in this work remains available in a public repository.

## 7 Acknowledgment

We would like to thank the Infodengue project (https://info.dengue.mat.br/), Brazil, for their contribution to data collection. We would also like to thank the Foz do Iguaçu Health Department and the city’s Zoonosis Control Center for making all data sets available and also for their valuable contribution to the interpretation and systematization of the collected data.

## 8 Author contribution statement

CC, STRP and FG conceived the research, RML contributed with data interpretation and the method design, CR, EA and FC developed the software. CR, EA, LB, FC and FG carried out the analyses and conducted the simulations, AL, CM, systematized epidemiological, entomological and climatic databases. CC and STRP acted as guarantors. All authors contributed to writing and reviewing the manuscript.

